# Peripheral immune profiles separate disease activity stages in Birdshot Uveitis

**DOI:** 10.64898/2026.05.27.26354201

**Authors:** D Pohlmann, I Kaeferstein, B Kruse, S Winterhalter, A Thiel, U Pleyer, J Braun

## Abstract

**Purpose:** To characterize peripheral immune alterations in treated birdshot uveitis (BU) patients using high-dimensional mass cytometry and multiplex serology.

**Design:** Cohort study.

**Subjects:** 36 BU patients on immunomodulatory treatment (IMT) and 31 healthy controls (HCs).

**Methods:** Detailed ophthalmologic examinations were performed, and peripheral blood and serum samples were collected for immune profiling using mass cytometry and multiplex cytokine analysis.

**Main Outcome Measures:** Imaging-based indicators of ocular inflammation; peripheral immune cell frequencies; serum cytokine levels.

**Results:** Compared to HCs, BU patients showed increased frequencies of Th17, CD146^+^ T cells, intermediate effector/central memory T cells co-expressing CXCR3 and CCR4, CD56^dim^ NK cells and elevated IL-18 levels. Patients were clinically stratified by an expert ophthalmologist into three disease activity groups: Inactive, Active (comprising combinations of surface retina, deep retina and choroid activity) and Burned-out. Inactive patients harbored more quiescent effector T cells, e.g. Tim-3^+^ Tc17-Tc22 intermediates and more CD8^+^ T_SCM_, potentially representing a resting pool of autoimmune T cells. Active patients exhibited increased *in vivo* activation of relevant T cells, with stronger HLA-DR, CD38 or PD-1 expression, and highest levels of CD56^dim^ NK cells.

Immune profiles were also linked to treatment subgroups: csDMARDs (conventional synthetic disease-modifying antirheumatic drugs) were associated with higher CD56^bright^ NK frequencies, and absence of therapy showed elevated PD-1⁺/SLAMF7⁺ Tc17+1 and PD- 1⁺CD57⁺ CD8⁺ T_EMRA_ cells. IL-6R blockade (tocilizumab) resulted in loss of IL-6R⁺ T-cells accompanied by increased SLAMF7⁺ T cells, due to epitope masking.

**Conclusions:** Peripheral CyTOF profiling anchored to thorough clinical stratification revealed disease activity-associated immune signatures and therapy-associated imprints in BU.

## INTRODUCTION

Birdshot Uveitis (BU), also known as Birdshot Chorioretinopathy (BCR), represents a rare, non-infectious posterior uveitis, characterized by recurrent episodes of inflammation affecting the retina and stromal choroid with “shotgun pattern” appearance, ovoid lesions around the optic nerve and mid-periphery of the retina^1^. An early clinical characteristic is retinal inflammation, marked by the leakage of retinal vessels, alongside inflammation of the choroidal stroma, manifested as hypofluorescent spots (dark dots) on imaging. Clinically, patients commonly experience floaters, blurred vision, and photopsia, and may encounter visual impairment encompassing reduced visual acuity (VA), nocturnal blindness and color blindness, resulting from retinal and macular degeneration, extensive choroidal depigmentation, and progressive formation of scar tissue on the retina. Histological studies have revealed severe retinal degeneration, near-complete loss of choroidal layers, and damage to the retinal pigment epithelium (RPE) in end-stage BU (burned-out patients)^2^. Our own previous data have confirmed irreversible retinal and choroidal damage in a subset of BU patients^3^.

Although BU is well-characterized clinically, its etiology and pathogenesis remain elusive. BU is highly associated with the major histocompatibility complex (MHC) gene human leukocyte antigen class I allele A*29 (HLA-A*29) and a variant of the Endoplasmatic reticulum aminopeptidase 2 (ERAP2) gene. These findings further support the classification of BU as an autoimmune disease and also a prototypic member of the recently proposed family of “MHC-I-opathies” ^4–9^. ERAP-1 and ERAP-2 are enzymes involved in antigen processing and presentation, trimming peptides to an optimal size for binding to MHC-I molecules^10–12^. This process is crucial for antigen presentation in T-cell mediated immunity. The combination of differential peptide processing by ERAP2 and the presence of HLA-A*29 may be pivotal for presentation of disease-associated self-peptides and molecular mimicker antigens, which break central T cell tolerance and initiate autoimmunity. The common model for autoreactive T cell priming supports a gut–immune axis and ocular recruitment on a “second hit”- hypothesis^13,14^.

According to previous studies, circulating and tissue-infiltrating CD146⁺ T cells, including Th1, Th/c17, and NK cell imbalance may orchestrate local inflammation, blood-retinal-barrier (BRB) breakdown and irreversible tissue damage. The adhesion marker CD146 (Melanoma Cell Adhesion Molecule, MCAM) mediates endothelial adhesion and migration, and is expressed on endothelial cells, also in the choriocapillaris, and subsets of infiltrating T cells ^15–18^. Differential leukocyte migration across the inner and outer BRB contributes to the spatial- temporal component of Birdshot lesions and the progressive and relapsing course of BU. Given the autoimmune nature of BU, it is necessary to dissect involved immune cells in greater detail to identify disease-specific immune signatures, also to better understand disease etiology. In this study, we applied high-dimensional mass cytometry and multiplex cytokine profiling to comprehensively characterize peripheral immune signatures in BU. Our goal is to identify disease-specific immune alterations to find cues about disease etiology and potential biomarkers that may enhance diagnostic precision and guide individualized immunotherapy.

## METHODS

### Patients and study design

This single-center, prospective study was performed in accordance with the Declaration of Helsinki and approved by the local ethics committee (Ethikkommission der Charité-Universitätsmedizin Berlin) (EA2/066/19). Prior to participating in the study, written informed consent was obtained from each patient. The study aimed to investigate a total of 36 patients with BU, affecting 72 eyes, along with a control group compromising 31 healthy controls (HC). The diagnosis of BU was made according to the research criteria of the international consensus conference (Levinson 2006)^19^. Additionally, all BU patients had been previously screened for positivity of the HLA-A*29:02 allele. HCs were not HLA-typed.

### Clinical assessment

On the clinical examination day, a comprehensive evaluation of each BU patient included the following assessments: 1) Visual acuity (VA) of each patient was assessed using the Early Treatment Diabetic Retinopathy Study (ETDRS) chart in decimal notation and converted into logarithm of minimum angle resolution (logMAR). 2) Slit lamp Examination and Indirect Fundoscopy: The ocular inflammation was classified according to the Standardization of Uveitis Nomenclature (SUN) Working Group criteria^20^. Vitreous haze (VH) as the degree of cellular infiltration was graded in 0=no infiltration, +0.5=minimal, +1=mild, +2=moderate, and 3=severe. 3) Spectral-domain optical coherence tomography (SD-OCT): High-resolution imaging of both the macula and optic nerve was performed using SD-OCT (SPECTRALIS®, Heidelberg Engineering, Heidelberg, Germany). The macula assessment involved the average central retinal thickness (CRT) and identifying the presence of cystoid macular edema (CME). The external limiting membrane (ELM), the ellipsoid zone, and the complex of RPE and Bruch’s membrane as well as the presence of an epiretinal membrane (ERM) was assessed. Furthermore, the optic nerve head assessment included measurement of thickness of the retinal nerve fiber layer (RNFL). The average RNFL can vary among individuals and is typically around 90 to 110 µm in the peripapillary region^21^. Swelling and thinning of the RNFL were recorded as additional information in the context of inflammation or degeneration. 4) Fundus autofluorescence (FAF) was performed with BluePeak™-blue laser and infrared reflectance by SD-OCT (SPECTRALIS®, Heidelberg Engineering, Heidelberg,Germany). Following fluorescent patterns were assessed: a) Healthy RPE cells typically show background level of autofluorescence; b) Areas with increased lipofuscin accumulation appear as brighter spots; c) Areas of RPE atrophy or damage may exhibit reduced or absent autofluorescence^22,23^. 5) Fluorescein angiography (FA) was performed after intravenous (IV) injection of 5 mL 20% sodium fluorescein to visualize retinal and macular leakage. 6) Indocyanine green angiography (ICGA) and FA were performed simultaneously on the SD-OCT (SPECTRALIS®, Heidelberg Engineering, Heidelberg, Germany). For ICGA, 500 mg ICGA (Cardiogreen; Peaselt, Lorei, Germany) diluted in 7.5 mL of 0.9% saline solution were administered IV. Findings from both FA and ICGA, were graded based on Tugal-Tutkun et al. to quantify the active intraocular inflammation at the posterior pole^24^, including hyperfluorescence of the optic disc, CME, staining of retinal blood vessels, and/or the presence of leakage of superficial vessels and deep capillary plexus. Macular leakage was assessed using the grading system introduced by Yannuzzi^25^. All assessments were based on observations made during the late phases of angiograms. Additionally, early-phase (<3 minutes), intermediate-phase (10 minutes), and late-phase (28–32 minutes) angiograms following ICGA injection were examined for signs of choroidal inflammation, such as the presence of hypofluorescent dots, fuzzy, and ill-defined choroidal vessels, and late-diffuse choroidal hyperfluorescence. Areas displaying atrophy were specifically excluded from consideration in these analyses. 7) Fundus Photography for the evaluation of choroidal lesions was obtained using Zeiss FF 450+ (Zeiss, Jena, Germany).

### Blood Sampling and cell isolation

40mL heparinized blood and 5mL serum were collected from each BU patient and HC for further analysis. *Peripheral Blood Monocular Cells (PBMCs)* were isolated from whole blood, mixed 1:1 with phosphate-buffered saline containing bovine serum albumin (PBS/BSA), by layering onto 10mL Biocoll per 40ml blood/PBS suspension (Merck) and centrifuged at 800xg for 25 minutes at room temperature (RT). The interphase was resuspended in 50mL of PBS/BSA and centrifuged at 310xg for 10 minutes at RT. After a second washing step the cell pellet was resuspended in 10mL PBS/BSA/EDTA. Cell counts were obtained using the CASY- cell counter and analyzer system (Roche Innovatis, Basel, Switzerland).

### Staining cells for mass cytometry

Up to 3×10^6^ PBMCs were transferred into individual wells and suspended in 2mL CyFACS (PBS / BSA / 2 mM EDTA/ 0.05% Natrium azide). After centrifugation at 310xg for 7 minutes at RT, the supernatant was removed. Antibody (Ab)-MasterMix was thawed and filtered through a 0.1µm centrifugation column. The cell pellet was incubated with 50µL of Ab- MasterMix at RT for 20 minutes. For dead cell labeling, samples were incubated with 5µM Cis- Platin for additional 10 minutes at RT.

The staining reaction was terminated by two washing steps with 2mL CyFACS buffer and centrifugation at 310xg for 7 minutes at RT. Subsequently, iridium intercalation was performed with Iridium intercalator solution with a final dilution of 1:5000 in MaxPar FixPerm buffer. The cell pellet was resuspended in 1mL of the intercalator solution and left to incubate for 60 minutes at RT. 1mL CyFACS was added, and centrifuged at 800xg for 7 minutes at RT. The supernatant was removed, and cells were resuspended in 250µL of the 1.6% PFA solution, incubating overnight at 4°C.

### Cryopreservation

The following day, antibody-labelled samples were washed with 2mL of CyFACS and centrifuged at 800xg for 7 minutes, and cell pellets were resuspended in 900µL FCS. Cell suspensions were transferred to cryotubes and placed in freezing containers (Mr. Frosty™, Thermo Scientific, Massachusetts, US), and 100µL DMSO added per cryotube. Finally, the cryotubes were moved to -80°C for overnight storage, followed by long-term storage in liquid nitrogen.

### Barcoding and measuring of stained and frozen PBMCs

Frozen and stained PBMCs were gently thawed at 37°C until nearly molten and transferred to 15mL Falcon tubes with 10mL Medium III (80% RPMI and 20% FCS). Subsequent centrifugation at 800xg for 10 minutes was performed, and the supernatant was carefully aspirated. The cell pellet was resuspended in 1mL Benzonase medium (Pierce Universal Nuclease, 25U/mL, 1:10,000 in Medium III) and incubated for 5 minutes at 37°C. The suspension was filled up to 10mL with CyFACS, centrifuged at 800xg for 10 minutes and aspiration of the supernatant.

MaxPar Barcode tubes were thawed for 10 minutes at RT before aliquoting 1mL MaxPar FixPerm buffer per individual sample. The contents of the barcode tubes were resuspended in 100µL MaxPar FixPerm buffer from the respective aliquots and mixed back into the original tubes. After incubation, the cell pellet was resuspended in 1mL Barcoding solution and incubated for 60 minutes at RT. The reaction was terminated by adding 1mL of CyFACS and centrifuged at 800xg for 7 minutes, followed by a second washing step with with 2mL CyFACS. Finally, cell pellets were resuspended in 200µL CyFACS and a sample volume of 2µL taken for pre-pool cell counting.

Samples were pooled to reach a final volume of 2mL in CyFACS and centrifuged at 800xg for 7 minutes. After supernatant aspiration, cell pellets were resuspended in 2mL water and centrifuged at 800xg for 7 minutes. Cell pellets were finally resuspended in 1mL of water. A 10µL sample volume was taken for cell counting. The pooled samples were divided into individual wells, each containing approximately 2×10^6^ cells after the final wash. Samples were brought up to a volume of 2ml with water and centrifuged at 800xg for 7 minutes. The supernatant was aspirated to completion and the cell pellet preserved on ice for subsequent measurements using CyTOF.

### Mass Cytometry Acquisition

We processed PBMCs of 36 BU patients and 31 HC using a high-dimensional 37-marker mass cytometry panel, with a primary focus on T cell phenotypes (see Supplementary table 1). To achieve a high-resolution dataset of 1 million events per sample, we pooled three samples (each containing approximantly 3.25 x 10^6^ cells), and acquired 3 million events on the CyTOF per pool, yielding an average of 8.67 x 10^5^ PBMCs per data file. With this comparably high event count, we were able to identify and quantify even minor T cell subsets with confidence.

### Multiplex cytokine assay

Cytokine and chemokine measurement from serum samples in 1:4 dilution was performed using a 20-plex bead array kit (Bio-Plex Pro Human Immunotherapy Panel, 20-Plex #12007975, Bio-Rad) according to manufacturer’s instructions using a Bio-Plex Pro Wash Station and measured on a Bio-Rad Luminex Bio-Plex 200 Multiplex ImmunoAssay Analyzer.

### Data/Statistical Analysis

The mass cytometric dataset was analyzed using two complementary approaches (Supp. Fig. 1). Firstly, unsupervised clustering via FlowSOM and UMAP identified 240 cell clusters across CD4⁺, CD8⁺ T cells, and NK cells (80 clusters per lineage) (Supp. Fig. 1). These were aggregated into 30 memory, 38 differentiation, or 73 unique phenotypic subsets (Supp. Fig. 1A-C). Secondly, manual gating focused on resolving rare T cell populations potentially missed by clustering. A comprehensive manual gating scheme retrieved 229 distinct T and 54 NK cell populations with a focus on differential chemokine receptor expression by memory T cells (Supp. Fig 1D, Supp.Fig. 2). In brief, memory T cell subsets were defined based on CD45RA and CCR7 expression: naïve (T_N_, CCR7^+^CD45RA^+^), central memory (T_CM_, CCR7^+^CD45RA^−^), effector memory cells (T_EM_, CCR7^−^CD45RA^−^), and terminally differentiated effector memory T cell re-expressing CD45RA (T_EMRA_, CCR7^−^CD45RA^+^). CD4^+^ and CD8^+^ memory T cells were further divided based on chemokine receptor expression (CCR6, CXCR3, CCR4, and CCR10) into helper and cytotoxic T cell (Th/Tc) subsets: Th1/Tc1, Th2/Tc2, Th17+1/Tc17+1, Th17/Tc17, and Th22/Tc22, as described by Loyal et al.^26^ In addition, CD56^+^ CD3^-^ cells were gated as NK cells, split into CD8^+/-^ CD16^+^ CD56^dim^ and CD8^+/-^ CD16^-^ CD56^bright^ NK cells. These subsets were further characterized regarding PD-1, CD57, Tim-3, HLA-DR and CD38 expression. Correlation analysis of comparable T cell populations obtained either from unsupervised clustering or manual gating demonstrated consistency between both analytic approaches (Supp. Fig 1E).

To delineate immune alterations specific to BU, we compared the immune profiles of BU patients to those of HCs using log₂ fold-change and false discovery rate (FDR) of Wilcoxon statistical comparisons (Fig. 3A).

Statistical analysis was carried out using R (version 4.5.1) and R-Studio (version 2025.05.1+513) to assess the relationship between clinical findings, imaging results, and experimental data. Statistical significance tests, including Wilcoxon test, ANOVA with Tukey’s post-hoc test or Pearson correlation were employed and results are displayed either as p- values or as asterisks *(*, p* ≤ *0.05; **, p* ≤ *0.01; ***, p* ≤ *0.001*).

## RESULTS

### Clinical Characteristics of the Patient Cohort

A total of 36 patients diagnosed with BU and 31 age-matched HCs were enrolled and investigated (Fig 1). All BU patients were HLA-A*29:02 positive. All included BU patients were not treatment-naïve and at recruitment received either no therapy or conventional synthetic disease-modifying antirheumatic drugs (csDMARDs; azathioprine, mycophenolate mofetil, or methotrexate), TNF-α-antagonists (infliximab or adalimumab), IL-6R-antagonist (tocilizumab), Ciclosporin A (CsA), or low-dose prednisolone (<10 mg/day). Detailed characteristics of anthropometrics, treatment regimens, clinical, and ophthalmologic status are listed in Table 1.

**Figure 1.**
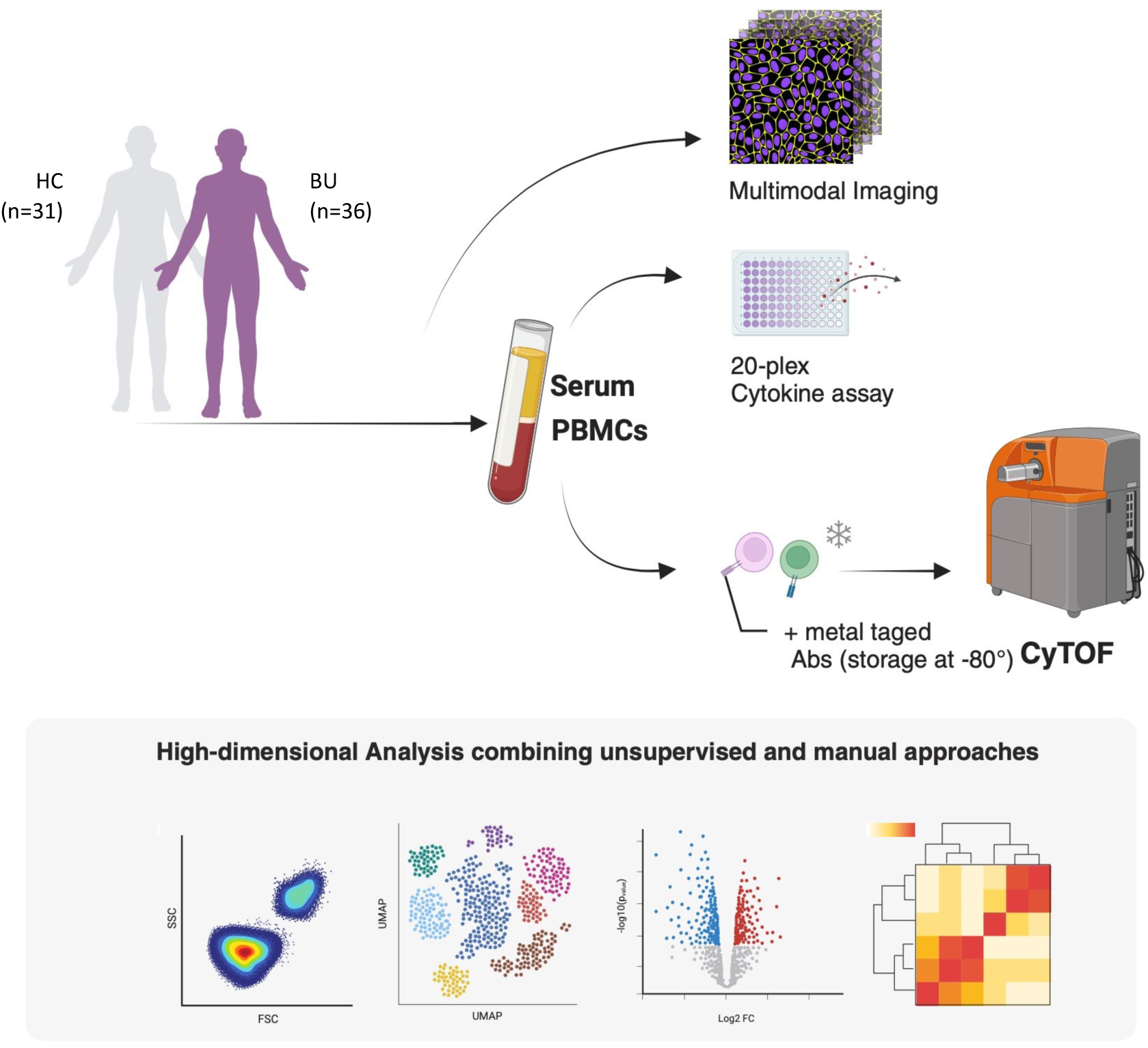
Study overview. Peripheral blood and serum were collected from birdshot uveitis (BU) patients (n=36) and healthy controls (HC, n=31). Multimodal ocular imaging was performed and PBMCs were stained with metal-tagged antibodies and acquired by mass cytometry (CyTOF). In addition, serum samples were analyzed by a 20-plex cytokine assay. High-dimensional analyses combined complementary pipelines — unsupervised clustering/UMAP, and targeted manual gating and differential abundance testing (log₂ fold-change/FDR).

**Table 1:**
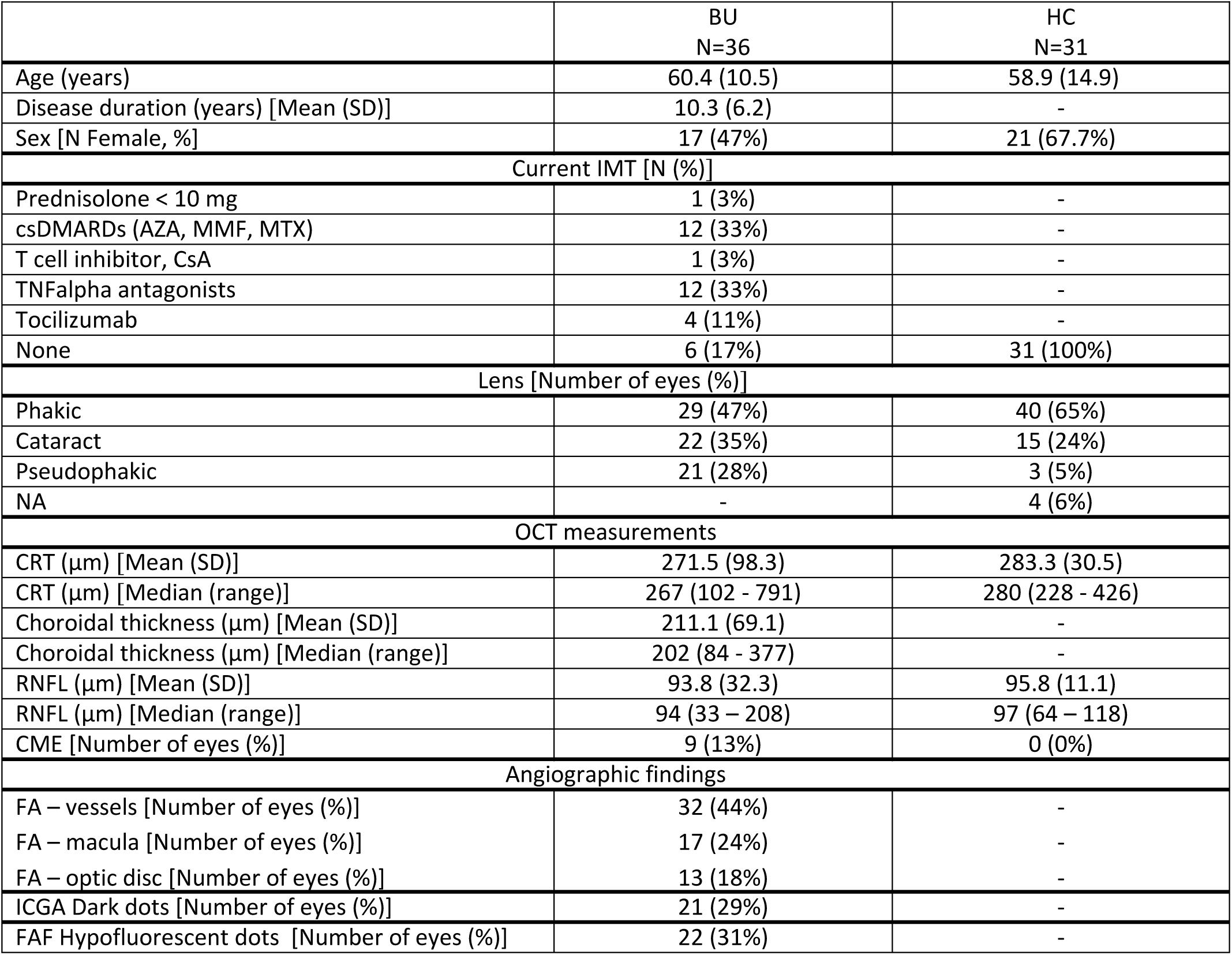
Baseline characteristics. OCT, Optical coherence tomography; IMT, Immunosuppressive treatment; AZA, Azathioprin; MTX, Methotrexate; CsA, Ciclosporin A; MMF, Mycophenolate mofetil; FAF, Fundus Autofluorescence, RNFL, Retinal nerve fiber layer; FA, Fluorescein angiography; ICGA, Indocyanine green angiography; CRT, Central retinal thickness; CME, Cystoid macula edema; csDMARD, conventional synthetic disease-modifying antirheumatic drugs.

|  | BU<br>N=36 | HC<br>N=31 |
| --- | --- | --- |
| Age (years) | 60.4 (10.5) | 58.9 (14.9) |
| Disease duration (years) [Mean (SD)] | 10.3 (6.2) | - |
| Sex [N Female, %] | 17 (47%) | 21 (67.7%) |
| Current IMT [N (%)] |  |  |
| Prednisolone < 10 mg | 1 (3%) | - |
| csDMARDs (AZA, MMF, MTX) | 12 (33%) | - |
| T cell inhibitor, CsA | 1 (3%) | - |
| TNFalpha antagonists | 12 (33%) | - |
| Tocilizumab | 4 (11%) | - |
| None | 6 (17%) | 31 (100%) |
| Lens [Number of eyes (%)] |  |  |
| Phakic | 29 (47%) | 40 (65%) |
| Cataract | 22 (35%) | 15 (24%) |
| Pseudophakic | 21 (28%) | 3 (5%) |
| NA | - | 4 (6%) |
| OCT measurements |  |  |
| CRT (μm) [Mean (SD)] | 271.5 (98.3) | 283.3 (30.5) |
| CRT (μm) [Median (range)] | 267 (102 - 791) | 280 (228 - 426) |
| Choroidal thickness (μm) [Mean (SD)] | 211.1 (69.1) | - |
| Choroidal thickness (μm) [Median (range)] | 202 (84 - 377) | - |
| RNFL (μm) [Mean (SD)] | 93.8 (32.3) | 95.8 (11.1) |
| RNFL (μm) [Median (range)] | 94 (33 – 208) | 97 (64 – 118) |
| CME [Number of eyes (%)] | 9 (13%) | 0 (0%) |
| Angiographic findings |  |  |
| FA – vessels [Number of eyes (%)] | 32 (44%) | - |
| FA – macula [Number of eyes (%)] | 17 (24%) | - |
| FA – optic disc [Number of eyes (%)] | 13 (18%) | - |
| ICGA Dark dots [Number of eyes (%)] | 21 (29%) | - |
| FAF Hypofluorescent dots [Number of eyes (%)] | 22 (31%) | - |

### Patient stratification and therapeutic regimens

To illustrate differences in disease stages of BU patients, all clinical, ophthalmologic and multimodal imaging parameters, including OCT measurements, VH, VA, and findings from FAF, ICGA, and FA were summarized in a heatmap (Figure 2A). Based on FA measurements, the expert investigator characterized disease activity (active, inactive) in four categories, namely activity overall (Activity FA), activity in the superficial retina (Type I), deep retina (Type II) or the choroid (Type III). By integrating the ophthalmologic expert rating, we generated the fine-mapped activity score (Activity Expert) with seven stages. Most groups with active disease were too small for meaningful statistical comparisons (n<5). Thus, for the following analyses, we compared HC and BU patients, which were split according to Activity General into Inactive, Burned-Out and Active and statistically compared. We further display five different stages of retina and choroid activity according to Activity Expert. Combined consideration of these activity stages, disease duration, patient’s age and the current treatment can provide cues for understanding disease progression (Fig. 2B-E).

**Figure 2.**
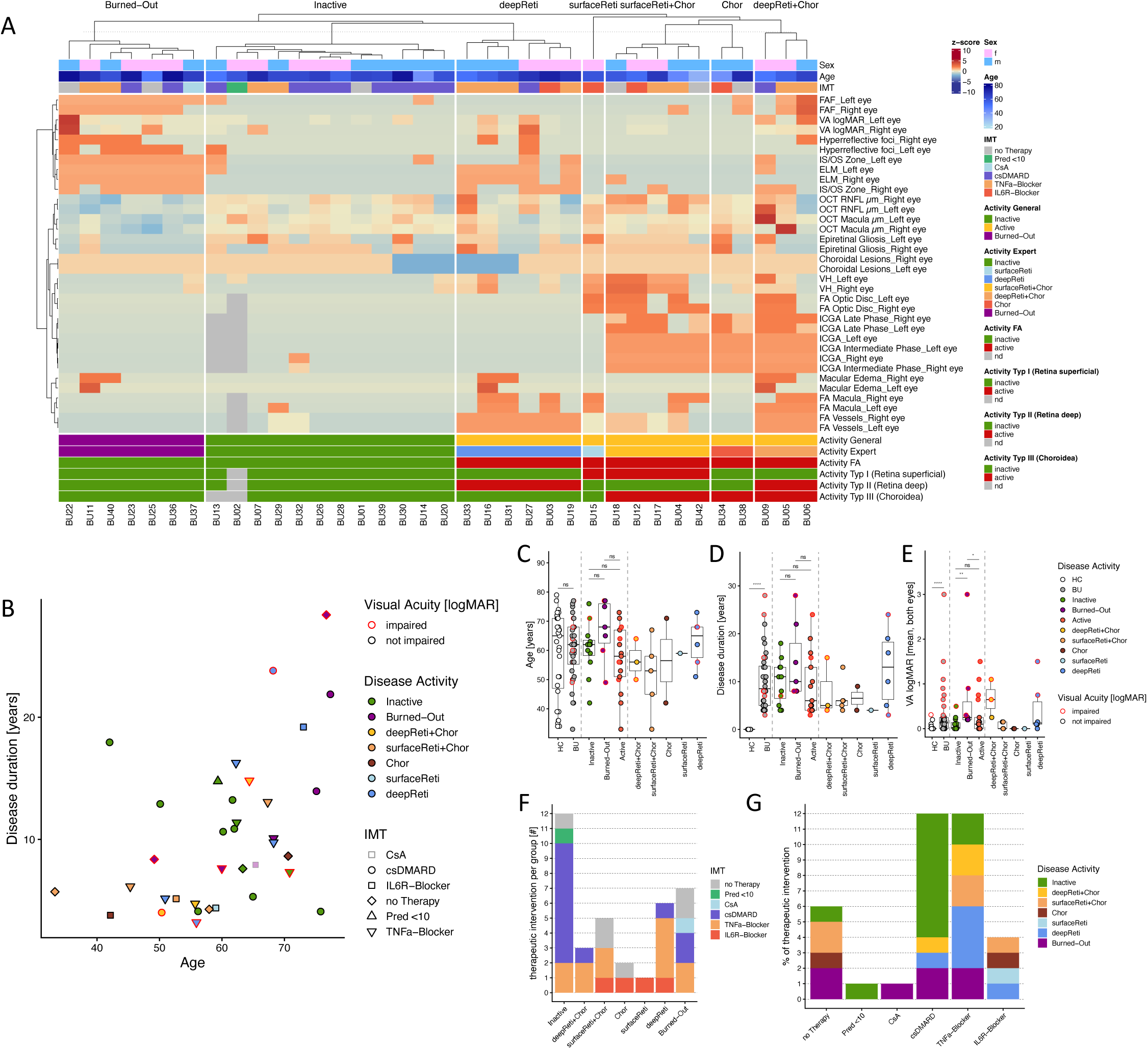
Multimodal clinical clustering of birdshot uveitis (BU) defines five disease activity groups, with accompanying age–disease duration distribution and treatment patterns across groups. **A**: Stratification of patients using key clinical and imaging variables. The heatmap shows clinical parameters split by expert-defined activity stages. Top annotations indicate age, sex, and current immunomodulatory treatment (IMT). Bottom annotation shows ophthalmologist-defined activity FA and different types (I = retina superficial; II = retina deep; III = choroid) based on fluorescence- and indocyaningreenangiography. These expert-defined activity gradings were summarised into Activity General with Inactive, Burned-Out and Active, which were further fine-mapped into surface Retina, surface Retina + Choroid, deep Retina, deep Retina + Choroid and Choroid (Activity Expert). **B**: Scatterplot of individual BU patients showing age versus disease duration. Points are color-coded by activity group and shaped by current immunosuppression; contouring in red indicates visual acuity (VA) impairment status. **C-E:** Box plots for age, disease duration and visual acuity with disease activity on the x-axis. Groups are HC and BU, split into Inactive, Burned-Out and Active, which were further split (indicated by dashed lines) according to activity affecting deep retina + chroroid (deepReti+Chor), surface retina + choroid (surfaceReti+Chor), choroid (Chor), surface and deep retina (surfaceReti and deepReti); for colors see legend. Horizontal bars and boxes indicate medians and interquartile ranges, respectively. Groups HC, BU, Inactive, Burned-Out and Active were statistically compared using Wilcoxon test. Statistical comparisons were performed using Wilcoxon test (*, p<0.05; **, p<0.01; ***, p<0.001 **F-G**: Treatment distribution by activity groups (F) and activity distribution by treatment (G) is displayed as stacked bar chart showing the number of current immunosuppressive regimens (IMT) within each group (Healthy control (HC), Inactive, Retina Activity, Retina + Choroid Activity, Choroid Activity, Burned-out).

Younger BU patients below 60 years had shorter disease duration and exhibited activity predominantly in both, retina (deep or surface) and choroid (surfaceReti+Chor; n=5 or deepReti+Chor, n=3), or just in the choroid (Chor, n=2). This was indicated by both FA and ICGA findings and the highest median VH, or by dark dots in the intermediate phase ICGA and sparse FA-leakage. Isolated retina activity (surfaceReti, n=1 and deepReti, n=6) was observed rather in older donors and was characterized by vascular leakage in the macular region and partially in the optic disc on FA imaging, without the presence of dark dots on ICGA. Inactive disease (n=12), characterized by absence of vascular leakage in FA and ICGA imaging, was mainly seen in patients of 55-70 years of age and disease duration of 5-15 years. Burned-out patients with severe visual impairment had the highest age (range 49-77) and longest disease duration (range 8-28 years). The burned-out stages exhibited loss of integrity in the ELM and the inner segment/outer segment (IS/OS Zone), and the lowest foveal and peripapillary retinal thickness in OCT measurements. Impaired visual acuity was present at highest rates in patients with activity in deep Retina and Choroid (deepReti+Chor, n=2/3), followed by deepReti (n=2/6) as well as Burned-out (n=3/7), and Inactive patients (n=1/11) (Fig. 2E).

Treatment plans were individualized for each patient, and a considerable number of BU patients had previously undergone various IMT regimens to control disease activity. Low dose Prednisolone and Cyclosporin A were each only given one patient. Most patients were treated with csDMARDs (n=12) and were mainly Inactive (n=8/12), or with TNF-α-antagonists (n=12), which exhibited mainly retina activity (n=8/12; deepReti+Chor, surfaceReti+Chor, deepReti; Fig 2F+G). Choroid active (surfReti+Chor, Chor) and Burned-out patients showed the highest number of patients without current treatment. IL-6R antagonists were administered to four Active patients.

### Distinct immune signatures in BU

Next, we analyzed the peripheral immunological profile by mass cytometry and related the identified cellular differences to the clinical activity groups. In BU patients, frequencies of Th17 T_CM_ subsets were significantly elevated, which was independent of disease activity (Fig. 3B, Supp. Fig. 4), in line with the autoimmune etiology of BU and previous reports^27^. Among Th17, a higher frequency of CD57^bright^ was observed in BU patients and specifically in Active disease (Fig. 3C). CD57 is characteristic for T cells with reduced proliferative capacity and often associated with chronic immune activation^28,29^. Serum IL-18 was significantly upregulated in BU patients, and highest in Inactive (Fig. 3D). In BU patients, we observed more CD146⁺ cells among overall CD4^+^ memory T cells and Th22 cells, specifically more abundant in Inactive patients (Fig. 3E+F). CD146, MCAM is involved in endothelial homing and associated with autoimmune and neuroinflammatory diseases^30–33^. In line with our previous report^1,34^, Active (and Burned-Out) patients had more Th1, less Th2 (Supp. Fig. 4) and thus a higher Th1/Th2 ratio in comparison to Inactive (Fig. 3G). Additionally, effector memory CD4⁺ T cells with co- expression of Th1- and Th2-associated chemokine receptors (CXCR3 and CCR4) were enriched in BU patients, especially in Active disease (Fig. 3H). Finally, HCs displayed significantly higher frequencies of CD56^bright^CD16⁻ NK cells (Fig. 3I), a subset typically associated with regulatory and tissue-repair functions. In contrast, CD56^dim^ NK cells were not altered in BU patients in general but significantly reduced in Inactive and increased in Active disease (Fig. 3J). This altered CD56^dim^/CD56^bright^ NK cell ratio in BU patients confirms reports about a potential imbalance in cytotoxic versus regulatory NK cell responses^35,36^.

**Figure 3.**
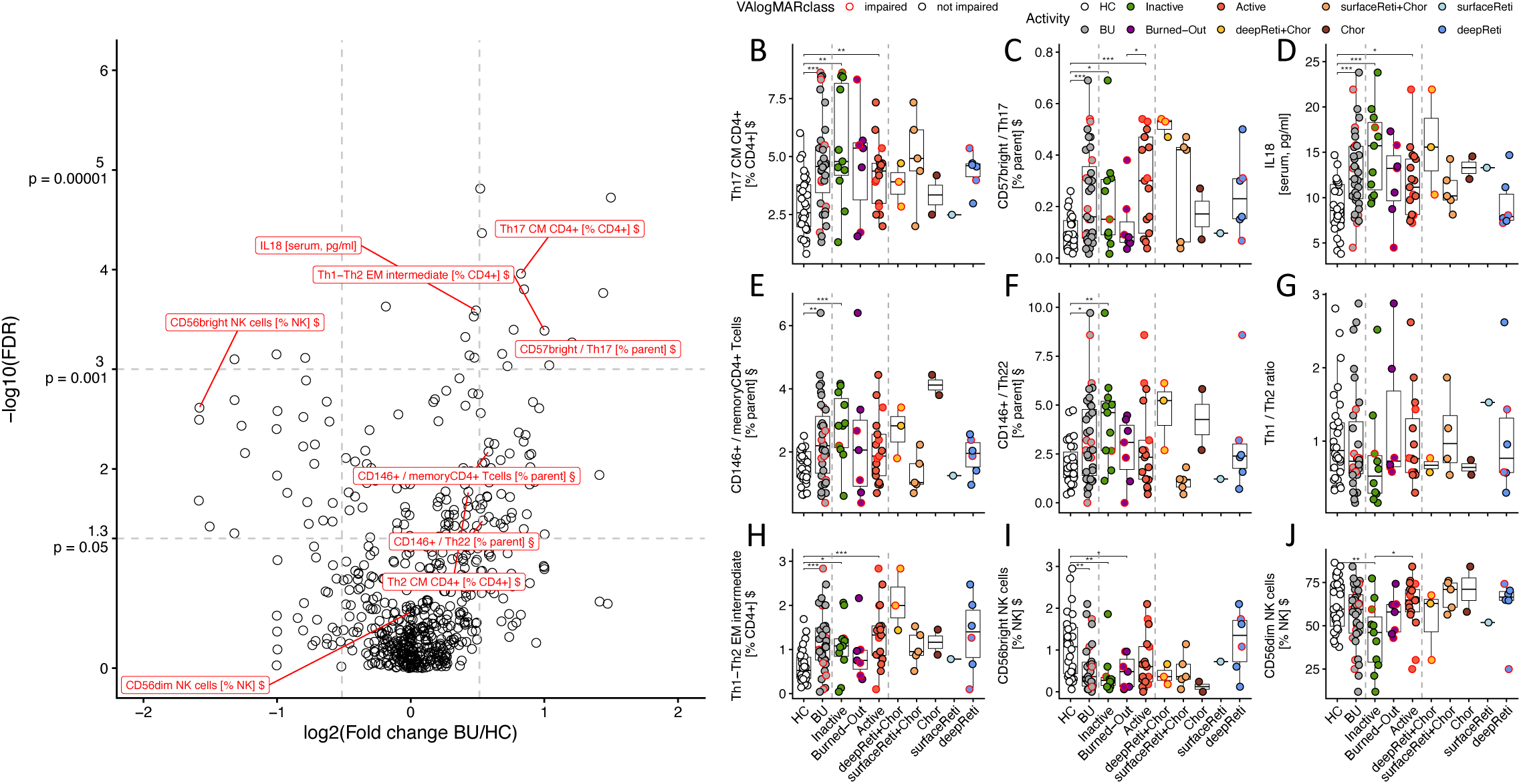
Peripheral immune signatures in birdshot uveitis (BU). **A:** Volcano plot of log₂ fold change (BU vs HC) versus –log₁₀(FDR) from CyTOF-derived immune cell subset frequencies. Labeled points denote the most significantly altered populations. Suffix “$” indicates subsets derived by FlowSOM. **B-G:** Box plots for representative parameters and subsets with disease activity on the x-axis. Groups are HC and BU, split into Inactive, Burned-Out and Active, which were further split (indicated by dashed lines) according to activity affecting deep retina + chroroid (deepReti+Chor), surface retina + choroid (surfaceReti+Chor), choroid (Chor), surface and deep retina (surfaceReti and deepReti); for colors see legend. Horizontal bars and boxes indicate medians and interquartile ranges, respectively. Groups HC, BU, Inactive, Burned-Out and Active were statistically compared using Wilcoxon test. Statistical comparisons were performed using Wilcoxon test (*, p<0.05; **, p<0.01; ***, p<0.001

### Disease activity-dependent immune signatures

To discover immune signatures associated with disease activity or therapy, we selected parameters correlating with Activity General and Activity Expert above a significance level of 0.05 and with Eta-Squared above 0.14 (Supp. Fig 5B) and clustered the medians per disease-activity group for the most relevant parameters (Fig. 4A). Accordingly, the Inactive group was characterized by higher levels of stem cell-like memory CD8^+^ T cells (T_SCM_) and more TIM-3^+^ among Tc17-Tc22 intermediate EM CD8^+^ T cells (Fig. 4B+C). T_SCM_ have been associated with autoimmunity, since they may represent a reservoir of autoimmunity-maintaining T cells, as shown for autoimmune uveitis, rheumatoid arthritis and SLE^37–39^. Especially the expansion of CD8^+^ T_SCM_ but reduction of CD4^+^ T_SCM_ seems to be a signature for autoimmunity. Tim-3^+^ Tc17- Tc22 may accumulate in Inactive because of chronic autoimmune activation. Tim-3 is an immune checkpoint inhibitor with reduced expression in several autoimmune diseases and polymorphisms described for inflammatory bowel disease, rheumatoid arthritis and others^40,41^. Active patients harbored more HLA-DR^+^ PD-1^+^ CD25^+^ Th17 and HLA-DR^+^ CD38^+^ Tc1 EM (Fig. 4D+E), while Burned-out patients displayed more PD1^+^ and CD57^+^ Tc1 EM cells (Fig. 4F+G), associated with exhaustion and senescence^42–44^. These observations clearly point at *in vivo* activation of autoimmunity-associated T cell subsets during Active disease stages and increased immune checkpoint expression in Inactive and Burned-out patients.

**Figure 4.**
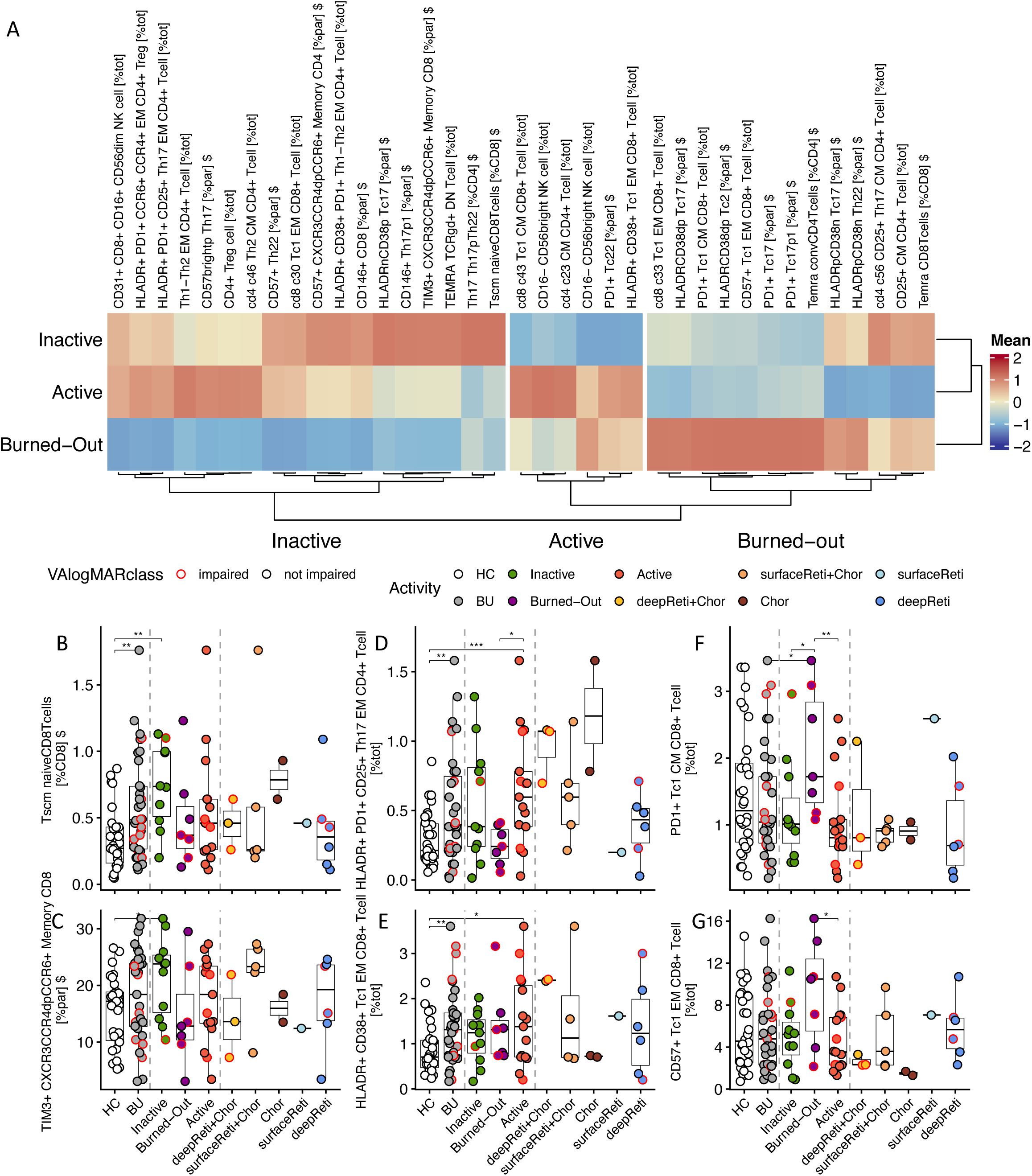
Disease activity–linked immune signatures delineate distinct clusters with shared features across groups. **A**: Heatmap of selected CyTOF subsets (rows) across groups (columns) showing median z-scores per group. Features were preselected by effect size (η² > 0.14) from ANOVA across activity states. **B-J**: Box plots for representative parameters and subsets with disease activity on the x-axis. Groups are HC and BU, split into Inactive, Burned-Out and Active, which were further split (indicated by dashed lines) according to activity affecting deep retina + chroroid (deepReti+Chor), surface retina + choroid (surfaceReti+Chor), choroid (Chor), surface and deep retina (surfaceReti and deepReti); for colors see legend. Horizontal bars and boxes indicate medians and interquartile ranges, respectively. Groups HC, BU, Inactive, Burned-Out and Active were statistically compared using Wilcoxon test. Statistical comparisons were performed using Wilcoxon test (*, p<0.05; **, p<0.01; ***, p<0.001

### Therapy-linked immune signatures

We next focused on immune profiles associated with current IMT, independent of clinical activity status (Supp. Fig. 5B), and compared the IMT group means of relevant parameters (Fig. 5A, Supp. Fig. 5).

**Figure 5.**
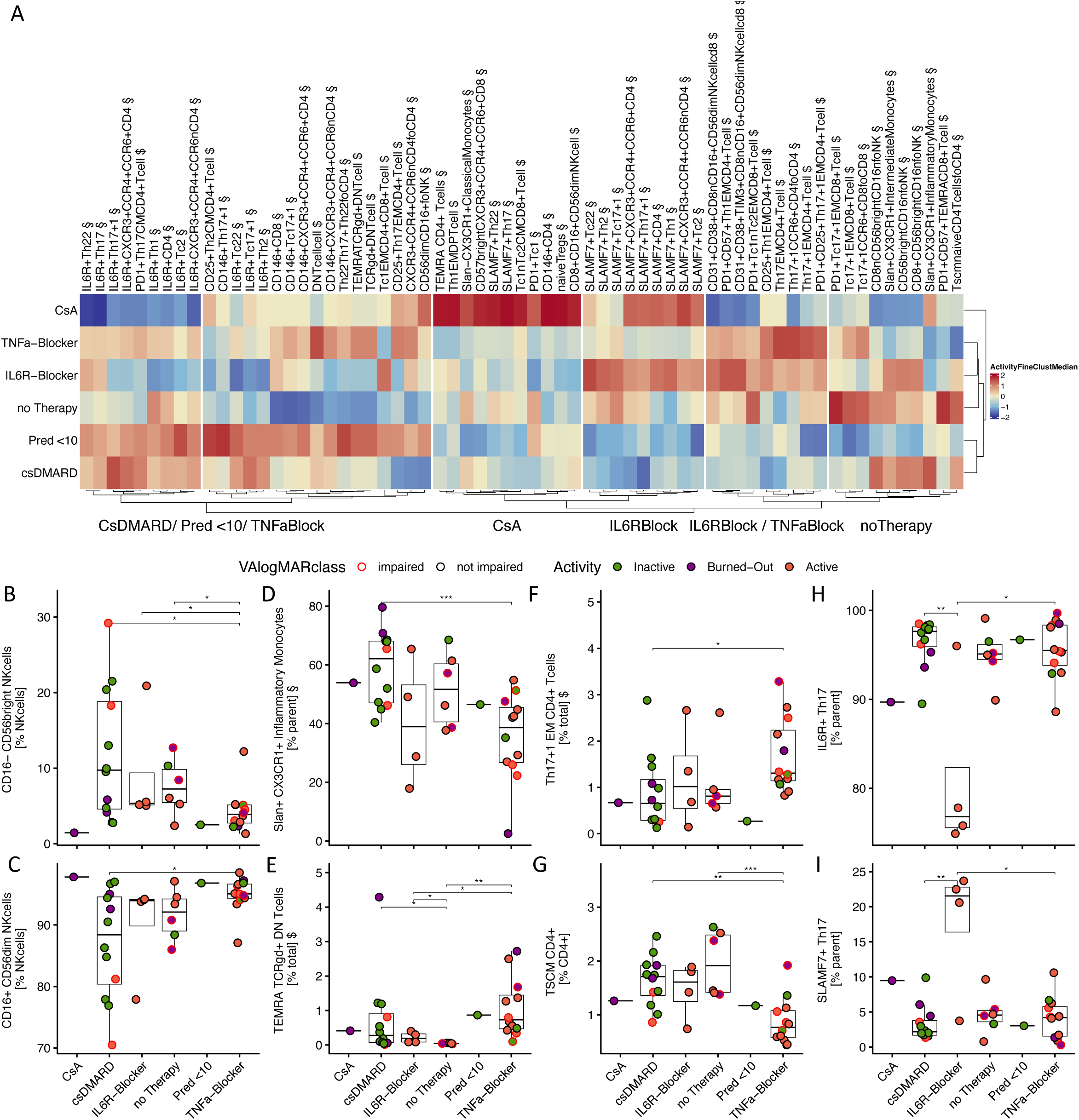
Therapy-linked immune signatures define distinct yet overlapping clusters. **A**: Heatmap of subsets (columns) stratified by current immunomodulatory therapy (rows: no therapy, csDMARDs, CsA, TNFα blocker, IL-6R blocker), showing median z-scores. **B-I**: Box plots for representative populations across therapy classes illustrate activity stage differences (e.g. SLAMF7⁺ and IL-6R+ Tc17+1, CD56dim/bright NK, inflammatory monocytes). Box plots show current IMT on the x-axis and dots are colored according to disease activity.

The csDMARD and TNF-α-antagonist group exhibit inverse immune profiles for many parameters. For example, CD56^bright^ CD16^−^ NK cells, Slan^+^ inflammatory monocytes and CD4^+^ T_SCM_ were highest for csDMARD and lowest for TNF-α-antagonist, while CD56^dim^ NK cells, TCRγδ^+^ T_EMRA_ and Th17+1 EM were low in csDMARD and high in TNF-α-antagonist treatment (Fig. 5A-G). The distinct group treated with the T cell inhibitor CsA, corresponding to a single patient, was characterized by markedly elevated frequencies of CD57^bright^ and CD146^+^ CD4^+^ T cell subsets (Fig. 5A), as well as a high proportion of CD56^dim^ CD16^+^ NK cells (Fig 5C). In the IL6R-antagonist-treated patients, a marked decline in IL-6R^+^ T helper subsets was accompanied by a reciprocal increase in SlamF7^+^ populations (Fig. 5H+I). Patients without current IMT had higher frequencies of PD-1⁺ and SLAMF7⁺ Tc17+1 cells, inflammatory monocytes, and PD-1⁺CD57⁺ CD8⁺ T_EMRA_ cells. However, it is difficult to untangle whether these signatures are definitely the result of the treatments itself or associated with the heterogeneous distribution of disease activity stages in the treatment groups.

## Discussion

In this study, we have utilized mass cytometry to immunophenotype and quantify more than 200 subsets of T cells and NK cells in BU patients and HC. We combined these immunophenotyping data and multiplex serological readouts with disease activity staging based on expert ophthalmologic assessment by multimodal imaging technologies.

In BU patients, we observed increased Th17 CM, CD146^+^ memory CD4^+^ T cells and reduced CD56^bright^ NK cells on cellular level and IL-18 on cytokine level. Increased CD146⁺ Th/c22 subsets and IL-18 levels in the peripheral circulation in BU patients may indicate recruitment to and/or recirculation from inflamed tissue sites. In addition, we observed more CXCR3/CCR4 double-positive T cells, representing intermediates between for instance Th1 and Th2. CXCR3 indicates Th1 priming and guides migration along a gradient of Chemokine Ligand (CXCL)9/10/11^45,46^. The enrichment of CXCR3⁺CCR4⁺ T_EM_ cells resembles patterns seen in psoriasis, in which this intermediate phenotype marks tissue-homing CD8⁺ T cells associated with disease relapse, connecting our findings to another MHC-I-mediated pathology^47^. Together, these findings represent T cell-centric autoimmune signatures and confirm previous reports^48^.

Current IMT had a large impact on immune profiles. Patients receiving csDMARD were predominantly Inactive and showed expansion of CD56^bright^ CD16^-^ NK cells, associated with immunoregulatory properties. CD56^bright^ CD16^-^ NK account for a small portion of the NK cell compartment but their expansion has been described to be associated with treatment efficiency in a case study of five uveitis patients treated with daclizumab, an IL-2R antagonist^49^. NK cell-mediated immunoregulation of activated T cells during daclizumab therapy has been shown earlier in Multiple sclerosis patients, with expansion of CD56^bright^ NK cells correlating with treatment response^50^. By comparison, in patients treated with TNF-α- antagonist CD56^bright^ NK cells remained comparatively low, while inflammatory monocytes showed a marked decline. Monocytes are a major source of TNF-α, and antagonizing TNF-α through infliximab or adalimumab may effectively reduce inflammatory monocyte differentiation^51^. Tocilizumab, a novel BU treatment option, was effective in treating refractory BU, showing a reduction in retinal vasculitis and CRT, while the impact on choroidal lesions has been less significant^52–54^. One patient from our study achieved sustained reduction of clinical activity later during treatment (data not shown). Strikingly, Tocilizumab caused a notable decrease in IL-6R⁺ subsets, accompanied by an increase in SLAMF7-expressing cell. While downregulation and internalization of the IL-6R with treatment can be expected, such dramatic decline in IL-6R subsets can be rather explained by epitope masking. Apparently, Tocilizumab can interfere with the staining antibody at the IL-6R binding site. This finding highlights the technical difficulties in monitoring treatment with biologicals.

The herein applied disease activity staging allows to describe the following sequence of events in BU, as summarized in Figure 6. Early disease (< 5 years post diagnosis) presents as Active, initially affecting the choroid and the deep retina, and later (> 5-10 years post diagnosis) rather exclusively the retina. This sequence may be explained by different immune privileged states of choroid and retina, conveyed by the two BRBs in the posterior eye. The outer BRB, encompassing choriocapillaris, RPE, and Bruch’s membrane, functions as the more immunomodulatory and dynamic epithelial barrier, whereas the inner BRB is more tightly regulated and less permeable, comparable to the blood–brain barrier^55,56^. Active episodes alternate with Inactive phases, in a relapse-remission cycle typical for autoimmunity. In late stages (>10 years), patients predominantly exhibit an end-stage, burned-out phenotype with simultaneous involvement of retina and choroid with dense T-cell infiltrates and structural loss of photoreceptor/RPE layers, underscoring chronic, compartment-spanning inflammation^57,58^. From an immunological standpoint, Inactive phases appear to be characterized by significantly increased CD8^+^ T_SCM,_ serum IL-18, and a skew towards Th22 and Th/c17, which suggests endothelial involvement. IL-18, often produced by epithelial cells, can increase IL-17 and IL- 22 production in Th17/Th22 as well as CD146 expression^59,60^. IL-22 in turn can increase endothelial permeability and proliferation, potentially sustaining leakage and vascular remodeling. Active disease was characterized by elevated Th17, Tc1, and CD146⁺ Th22, and a pronounced upregulation of exhaustion markers like PD-1 and CD57, e.g. on HLA-DR⁺ PD- 1⁺ Th17 T_EM_ subsets. The PD-1 phenotype aligns with a state of chronic antigen exposure and epigenetically imprinted exhaustion in tissue-engaged T cells^61,62^. Concomitant CD57^bright^ marks terminal differentiation or senescence with reduced proliferative capacity but preserved cytotoxic potential, a feature typical for long-standing inflammation^63^. Mature CD56^dim^ NK cells increased in Active patients may interact with inflamed epithelial cells and the IL-15/IL-18 microenvironment as part of the barrier-tissue activation^64^. Burned-out patients, traditionally considered inactive, do not appear immunologically quiescent, but represent a late, chronically antigen-experienced state with persistent cytotoxic memory despite absent angiographic leakage. T cells exhibited clear signs of exhaustion, with PD-1 and CD57 expression. In our pilot study, we observed increased CD8⁺ T_EMRA_ in Inactive patients^65^. T_EMRA_ subsets accumulate with age, an effect that influenced the findings in our Burned-out cohort but could not fully account for the increased frequencies compared to HCs. While TCRγδ expression was not examined in the pilot study, both findings indicate that increased T_EMRA_ frequencies are indicative for advanced disease, even in angiographic inactive eyes^65^. These patterns support a working model for an etiologic sequence, in which choroidal inflammation and outer BRB disruption appear early in disease, followed by retinal involvement, and subsequent accumulation of exhausted/terminally differentiated T cell states. These peripheral immune readouts change with disease activity, which explains why cross-sectional blood studies have been inconsistent and underscores the value of our disease activity-aware design. In future BU trials, stratifying patients with Retina vs. Choroid Activity and incorporating blood biomarkers as secondary endpoints may increase power and interpretability. A compact flow cytometry panel enabling assessment of functionally different Th/Tc subsets and CXCR3⁺CCR4⁺ intermediates, their *in vivo* activation (HLA-DR, CD38) or exhaustion and senescence (PD-1/CD57/Tim-3), as well as the CD56^dim/bright^ NK cell ratio could complement multimodal imaging to characterize disease activity and detect subclinical shifts.

**Figure 6:**
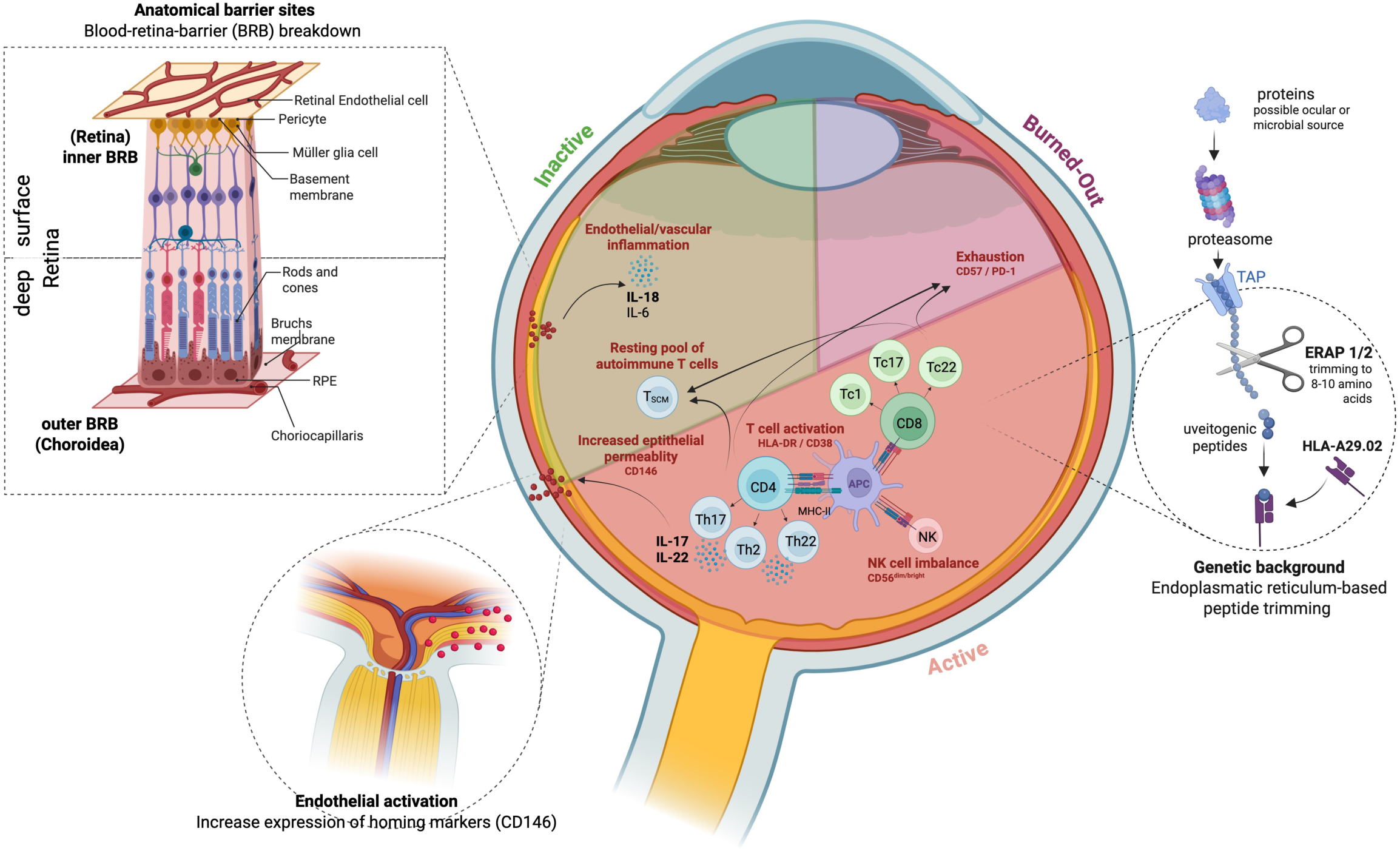
Schematic summary. HLA-A29 allele and ERAP variants 1 and 2 genetically determine the antigenic landscape in BU patients. T and NK cell activation and trafficking through the anatomical barrier sites (inner and outer blood retina barrier) differentially contribute to the different disease activity states and the progressing and relapse- prone nature of BU. Created with BioRender.

## Conclusion

In this study, we combined therapy-aware mass cytometry study in BU combining a clinically phenotyped cohort with multimodal imaging. Prior reports have profiled selected cytokines or limited lymphocyte compartments, but none integrated high-dimensional peripheral immune profiling and serology with clinical stratification. We were able to align compartment-specific activity with peripheral immune profiling. Our analysis did not include therapy-naïve patients but patients without current treatment, limiting conclusions about BU pathogenesis versus therapeutic effects. Limitations are inherent to peripheral sampling (intraocular events are inferred), modest sample sizes for certain disease activity and therapy subgroups, and a T/NK cell-focused panel that did not comprehensively profile myeloid/B-cell subsets. Using our dataset as a starting point for a standardized flow cytometry panel could support treatment monitoring and guide rational IMT switching for BU patients. Integration of antigen specificity and TCR clonality will be important next steps.

## Data Availability

All data produced in the present study are available upon reasonable request to the authors.

