## Supplemental Figures for "Peripheral immune profiles separate disease activity stages in Birdshot Uveitis"

### Supplementary Table 1
### Antibody Panel

| Isotope | Antigen | Manufacturer | CatalogID | Clone |
|---|---|---|---|---|
| 89Y | CD45 | Biolegend | 304045 | HI30 |
| 141Pr | CD1c | Biolegend | 331502 | L161 |
| 142Nd | CD57 | Biolegend | 322325 | HCD57 |
| 143Nd | TCRgd | Biolegend | 331202 | B1 |
| 144Nd | CD15 | Biolegend | 323035 | W6D3 |
| 145Nd | CD146 | Biolegend | 361002 | P1H12 |
| 146Nd | CCR4 / CD194 | Biolegend | 359402 | L291H4 |
| 147Sm | Slan | Miltenyi | 130-095-212 | DD-1 |
| 148Nd | CD69 | Biolegend | 310939 | FN50 |
| 149Sm | CD127 | Biolegend | 351337 | A019D5 |
| 150Nd | CD27 | Biolegend | 302839 | O323 |
| 151Eu | CD123 | Biolegend | 306027 | 6H6 |
| 152Sm | CD45RA | Biolegend | 304143 | HI100 |
| 153Eu | CX3CR1 | Biolegend | 341602 | 2A9-1 |
| 154Sm | TIM-3 | Biolegend | 345019 | F38-2E2 |
| 155Gd | CD14 | Biolegend | 301843 | M5E2 |
| 156Gd | CD183_CXCR3 | Biolegend | 353733 | G025H7 |
| 158Gd | CCR10 | Biolegend | 341502 | 6588-5 |
| 159Tb | PD-1 | Biolegend | 329941 | EH12.2H7 |
| 160Gd | CD28 | Biolegend | 302937 | CD28.2 |
| 161Dy | CD8 | In-house |  | GN11137 |
| 162Dy | IgD | Biolegend | 348235 | IA6-2 |
| 163Dy | CD56 | Biolegend | 318345 | HCD56 |
| 164Dy | CD95 | Biolegend | 305631 | DX2 |
| 165Ho | CCR6 / CD196 | Biolegend | 353427 | G034E3 |
| 166Er | CD19 | Biolegend | 302247 | HIB19 |
| 167Er | CCR7 | Biolegend | 353237 | G043H7 |
| 168Er | CD4 | In-house |  | TT1 |
| 169Tm | CD25 | Biolegend | 356102 | M-A251 |
| 170Er | CD3 | BD | 555330 | UCHT1 |
| 171Yb | SLAMF7 / CD319 | Biolegend | 331802 | 162.1 |
| 172Yb | CD38 | Biolegend | 303535 | HIT2 |
| 173Yb | CD141 | Biolegend | 344102 | M80 |
| 174Yb | CD31 | Biolegend | 303127 | WM59 |
| 175Lu | CD126_IL6R | Biolegend | 352802 | UV4 |
| 176Yb | HLA-DR | Biolegend | 307651 | L243 |
| 209Bi | CD16 | Biolegend | 302051 | 3G8 |

### Supplementary Figure 1

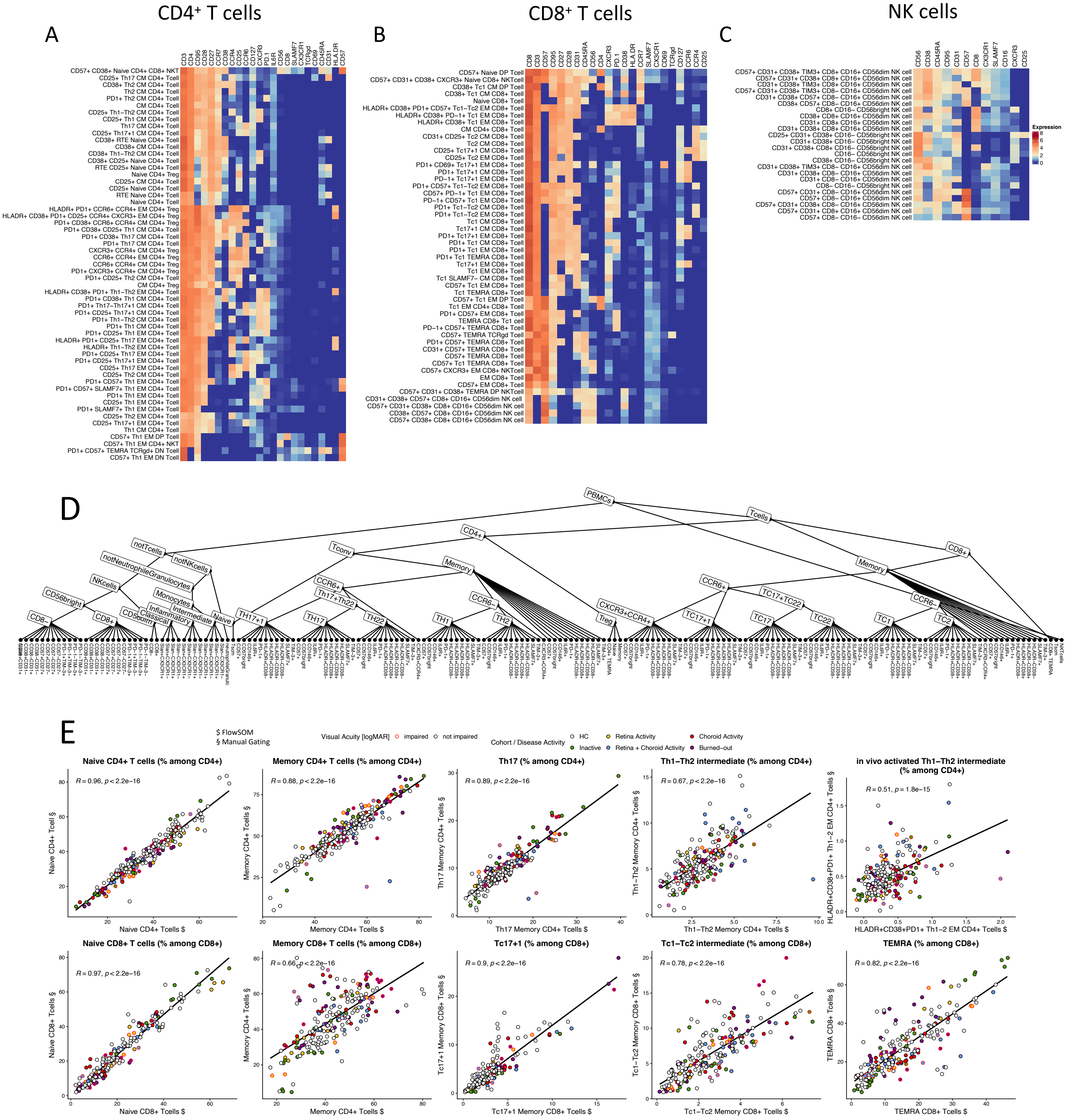


**Supplementary Figure 1. Masscytometry immunoprofiling of T and NK cells using FlowSOM and manual gating confirming comparable results and focusing on rare T cell populations.**

**A-C:** CD8$^{+}$ and CD4$^{+}$ T cells as well as NK cells were clustered using FlowSOM (240 cell clusters), followed by manual cluster annotation and UMAP embedding. **D:** Hierarchical scheme of the T and NK cell-focused gating strategy. **E:** Correlation of FlowSOM (indicated as "$")- and manual gating(§)-derived cell frequencies of matching populations revealed high comparability of results. Pearson correlation coefficients R and p-values are displayed.

### Supplementary Figure 2

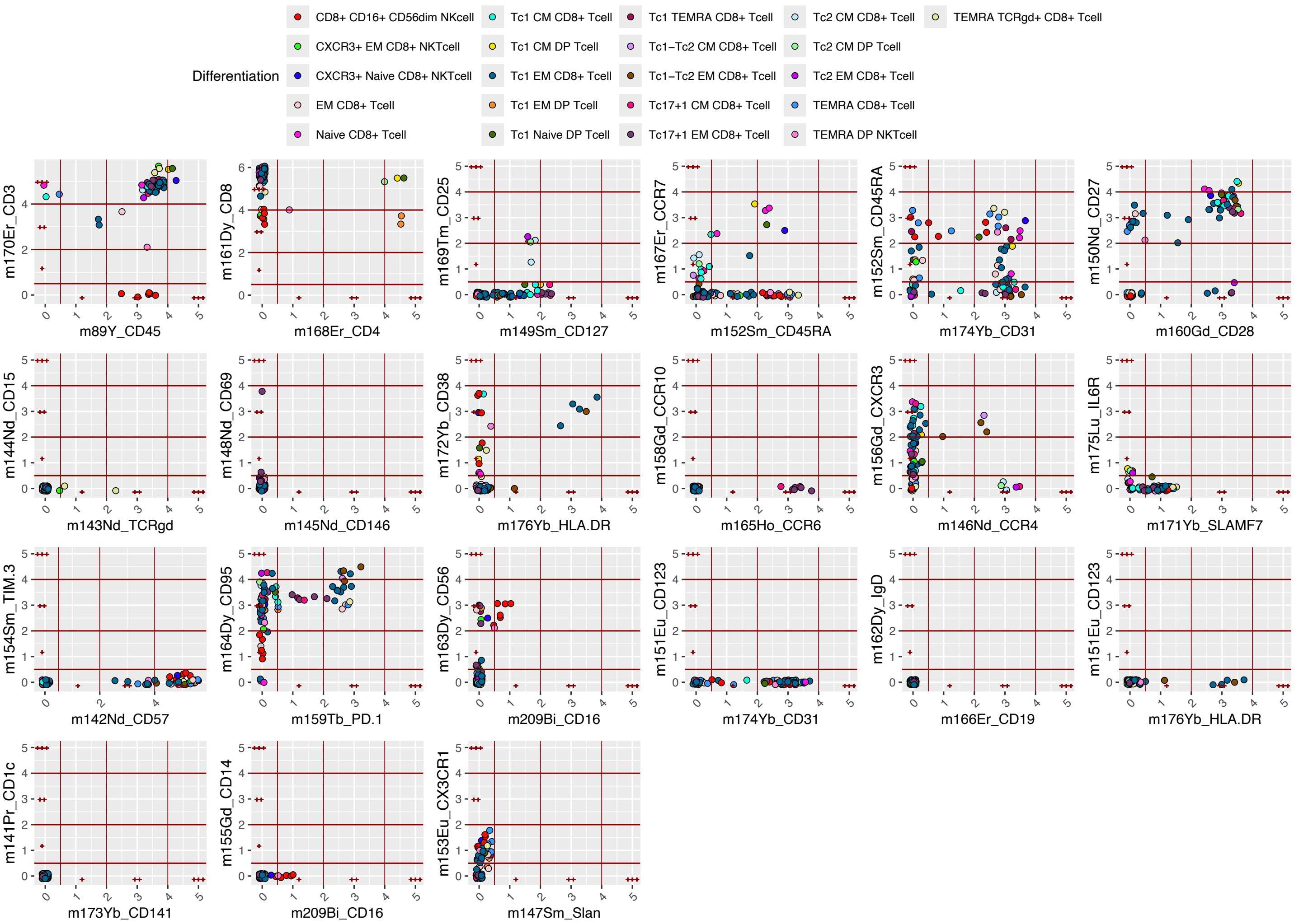


**Supplementary Figure 2: Phenotype annotation of FlowSOM-derived clusters.**
FlowSOM-derived clusters were assigned with phenotype annotations using means of marker expression. Here, exemplary differentiation phenotypes (such as Tc1, Tc17 etc.) among CD8 cells is shown. See Appendix for all phenotypes.

### Supplementary Figure 3

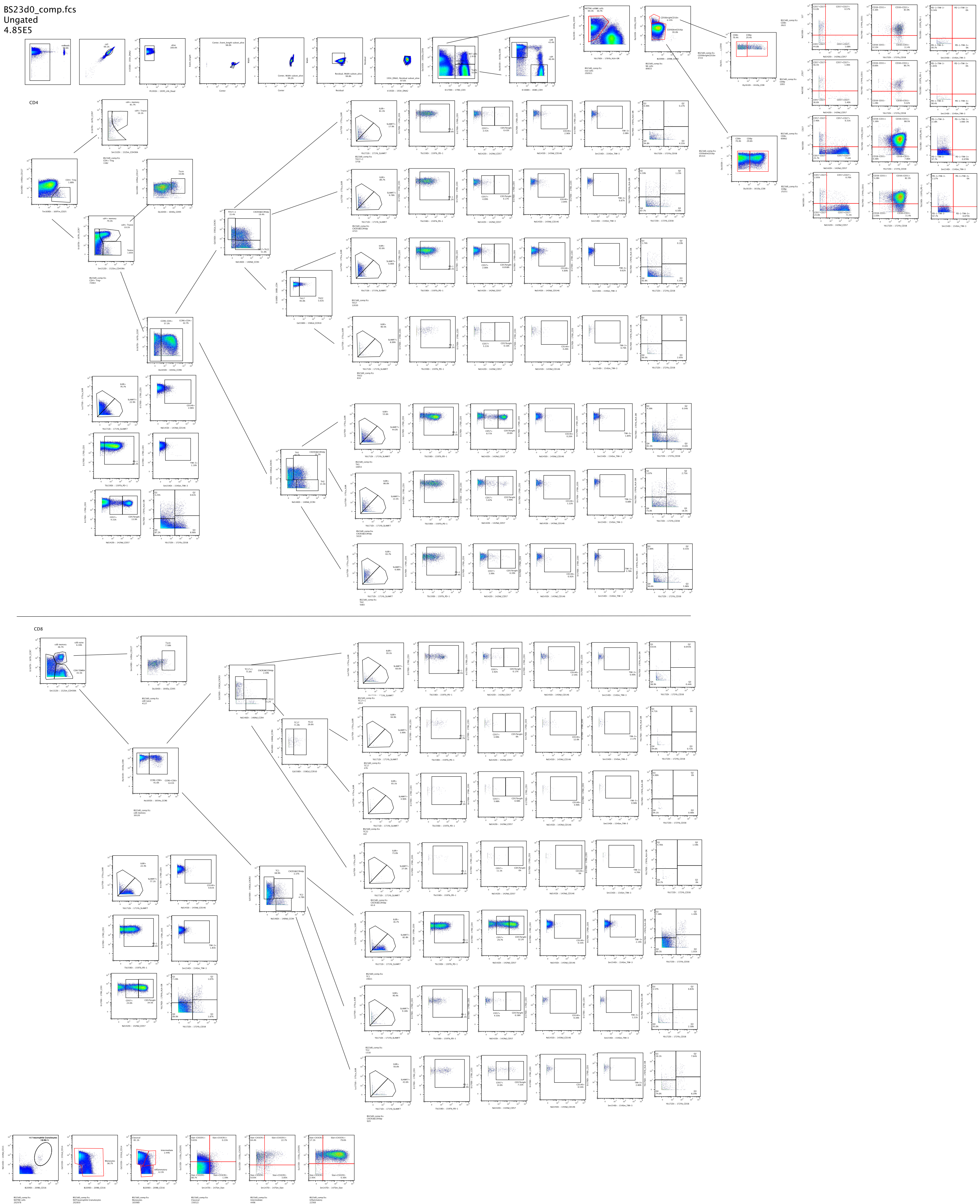


**Supplementary Figure 3: Manual gating.**

### Supplementary Figure 4

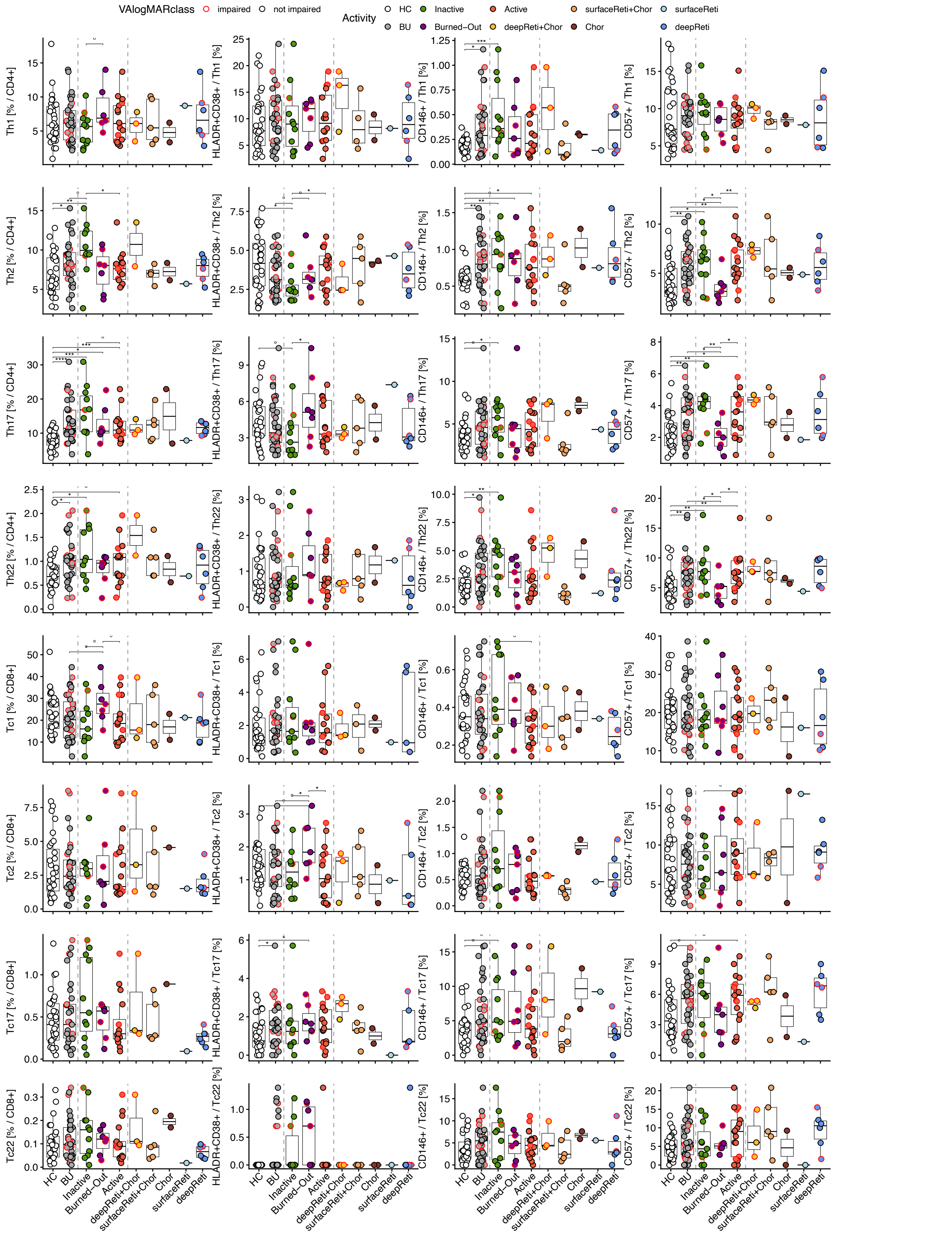


**Supplementary Figure 4: Overview about TH and TC subsets.**

**B-J**: Box plots for representative parameters and subsets with disease activity on the x-axis. Groups are HC and BU, split into Inactive, Burned-Out and Active, which were further split (indicated by dashed lines) according to activity affecting deep retina + chroroid (deepReti+Chor), surface retina + choroid (surfaceReti+Chor), choroid (Chor), surface and deep retina (surfaceReti and deepReti); for colors see legend. Horizontal bars and boxes indicate medians and interquartile ranges, respectively. Groups HC, BU, Inactive, Burned-Out and Active were statistically compared using Wilcoxon test. Statistical comparisons were performed using Wilcoxon test (*, p<0.05; **, p<0.01; ***, p<0.001

### Supplementary Figure 5

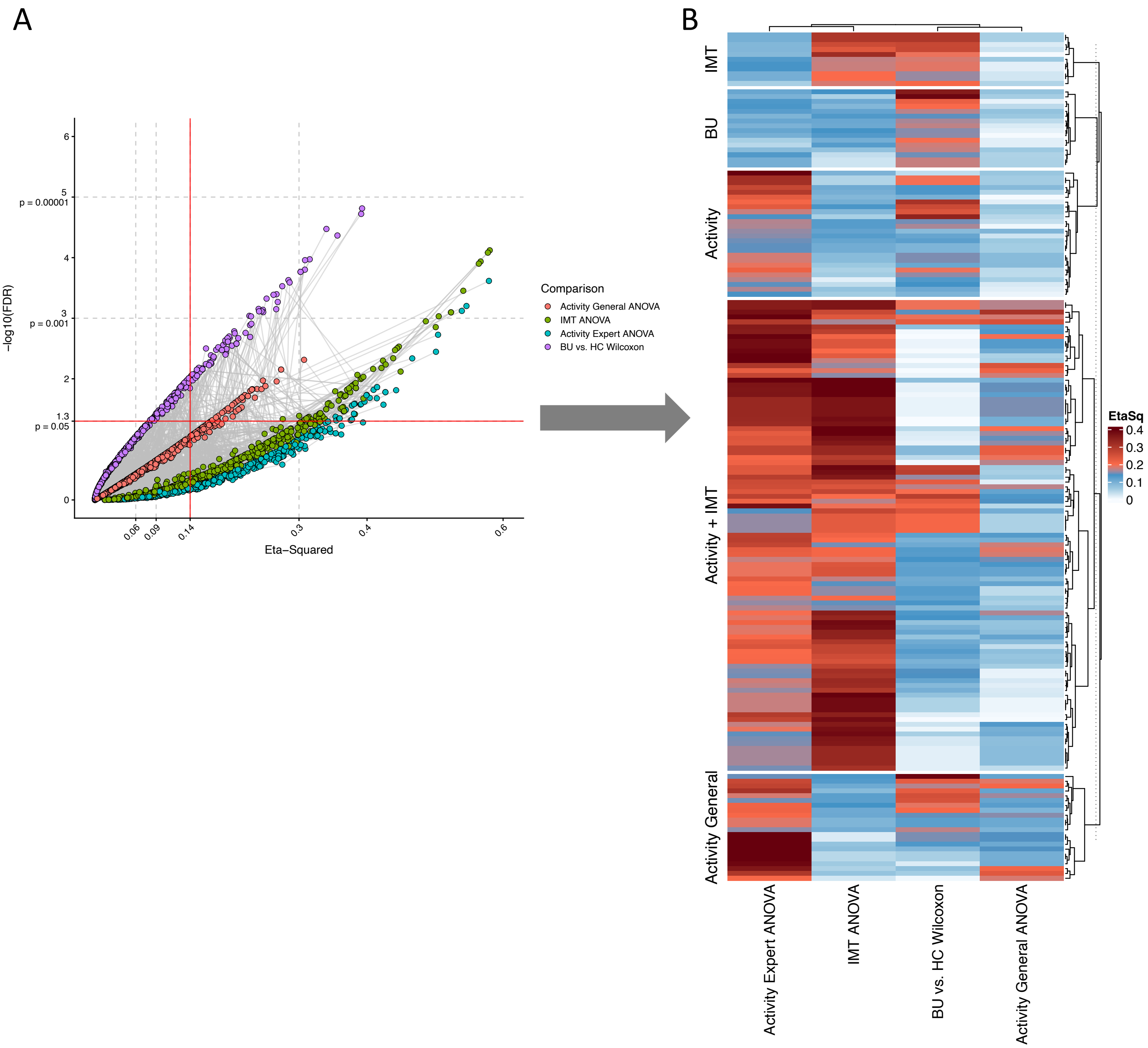


**Supp. Figure 5: Effect sizes of several comparisons allow identifification of parameters associated with BU alone, Activity, IMT or Activity and IMT.**

**A:** Volcano plot of Eta-Squared versus negative $log_{10}$(FDR) from Wilcoxon test for Cohort or Anova over different disease activity strata or treatment regiments (comparison indivated by color). **B:** Heatmap showing Eta-Squared across different comparisons of parameters filtered for significance in at least one comparison ($p <= 0.05$). Colors indicate Eta-Squared of indicated statistical test. Parameters with effect sizes greater 0.14 were grouped as indicated by the row group titles and further investigated separately in Figure 4 and 5.
